# Association of cumulative exposure to triglyceride and remnant cholesterol with the risk of cardiovascular disease in hypertensive patients with target LDL-C

**DOI:** 10.1101/2024.12.19.24319387

**Authors:** Shuang Guo, Chi Wang, Taoyu Hu, Lihua Lan, Zhen Ge, Jianxiang Huang, Shuohua Chen, Shouling Wu, Hao Xue

## Abstract

**Background:** The associations of cumulative exposure to triglycerides (cumTG) and remnant cholesterol (cumRC) with cardiovascular disease (CVD) risk in patients with optimal low-density lipoprotein cholesterol (LDL-C) levels remain uncertain. We aimed to explore the relationship between cumTG and cumRC and the risk of CVD in hypertensive patients with target LDL-C.

**Methods:** A total of 12,086 hypertensive participants with target LDL-C level who completed three health examinations from 2006 to 2010 were enrolled and followed until December 31, 2022. The cumulative burden was derived by multiplying the average level with the interval between two consecutive assessments. The primary outcome included a composite of CVD events (myocardial infarction, ischemic stroke, and hemorrhage stroke). Participants were divided into 4 groups based on the median values of cumTG and cumRC to assess their joint associations. Cox proportional hazard models were used to calculate the hazard ratio (HR) and 95% confidence intervals (CI) between the cumulative burden and CVD risk.

**Results:** During a median follow-up of 11.95 years, 1,392 participants developed CVD. After multivariable adjustment, compared to the low cumTG/low cumRC group, both the high cumTG/high cumRC group (HR 1.43, 95% CI 1.24‒1.64) and the low cumTG/high cumRC group (HR 1.26, 95% CI 1.07‒1.49), but not the high cumTG/low cumRC group (HR 1.01, 95% CI 0.84‒1.20), were associated with higher risk of CVD.

**Conclusions:** In hypertensive patients who met the target level of LDL-C, the cumulative burden of RC was a risk factor for CVD and exhibited a stronger association with CVD risk compared to cumulative triglycerides burden.

## Introduction

High concentrations of triglyceride-rich lipoproteins (TRLs) are recognized as a significant residual risk factor of atherosclerotic cardiovascular disease (ASCVD) when low-density lipoprotein cholesterol (LDL-C) reaches the target levels [1–2]. Given that triglyceride (TG) can be catabolized by most cells, but cholesterol cannot [3], remnant cholesterol (RC), the cholesterol content of TRLs, has been hypothesized to be the atherogenic component in TRLs. Evidence from genetic and epidemiological studies has demonstrated that elevated RC levels are associated with increased ASCVD risk independent of LDL-C levels [4–7].

The 2023 Chinese Lipid Management Guidelines recommend TG as a management target for patients with high-risk of ASCVD and target-levels of LDL-C [8]. As the content of TRLs and the risk factor of ASCVD, It is still a matter of debate whether elevated RC primarily contributes to the progression of ASCVD, rather than the TGs themselves. Furthermore, it remains unclear whether TG and RC independently predict ASCVD risk or exhibit a synergetic effect. Therefore, we conducted this study to investigate the separate and combined effects of TG and RC on ASCVD in hypertensive patients with target LDL-C levels. Since the visit-to-visit variability in lipid levels is associated with cardiovascular disease (CVD) [9], the cumulative burden of lipid exposure, an index that is potentially superior to a single measurement, has been recommended [10].

The present study aimed to examine the association between cumulative triglyceride (cumTG) and cumulative RC (cumRC) and the risk of CVD in hypertensive patients with target LDL-C levels based on the Kailuan cohort study.

## Methods

### Study design and Participants

The present study was based on the Kailuan Cohort Study, which was carried out from June 2006 to October 2007 and included 101,510 participants in the Kailuan community in Tangshan, China. Participants were followed biennially with repeated questionnaires and medical examinations. The detailed design of the Kailuan Cohort Study has been described previously [11].

In the current study, we included hypertensive patients (blood pressure ≥140/90 mm Hg, current use of antihypertensive therapy, or physician diagnosis of hypertension [12]) who attended health examinations in 2006/2007, 2008/2009, and 2010/2011. The exclusion criteria were as follows: (1) participants with missing lipid data; (2) participants whose LDL-C levels did not meet the target criteria, defined as <3.40 mmol/L for low-risk patients and <2.60 mmol/L for moderate-to-high-risk patients, as specified by the 2023 Chinese Lipid Management Guidelines [8]; (3) participants with a history of CVD. Finally, a total of 12,086 participants were included in the final analyses (Figure 1).

**Figure 1.**
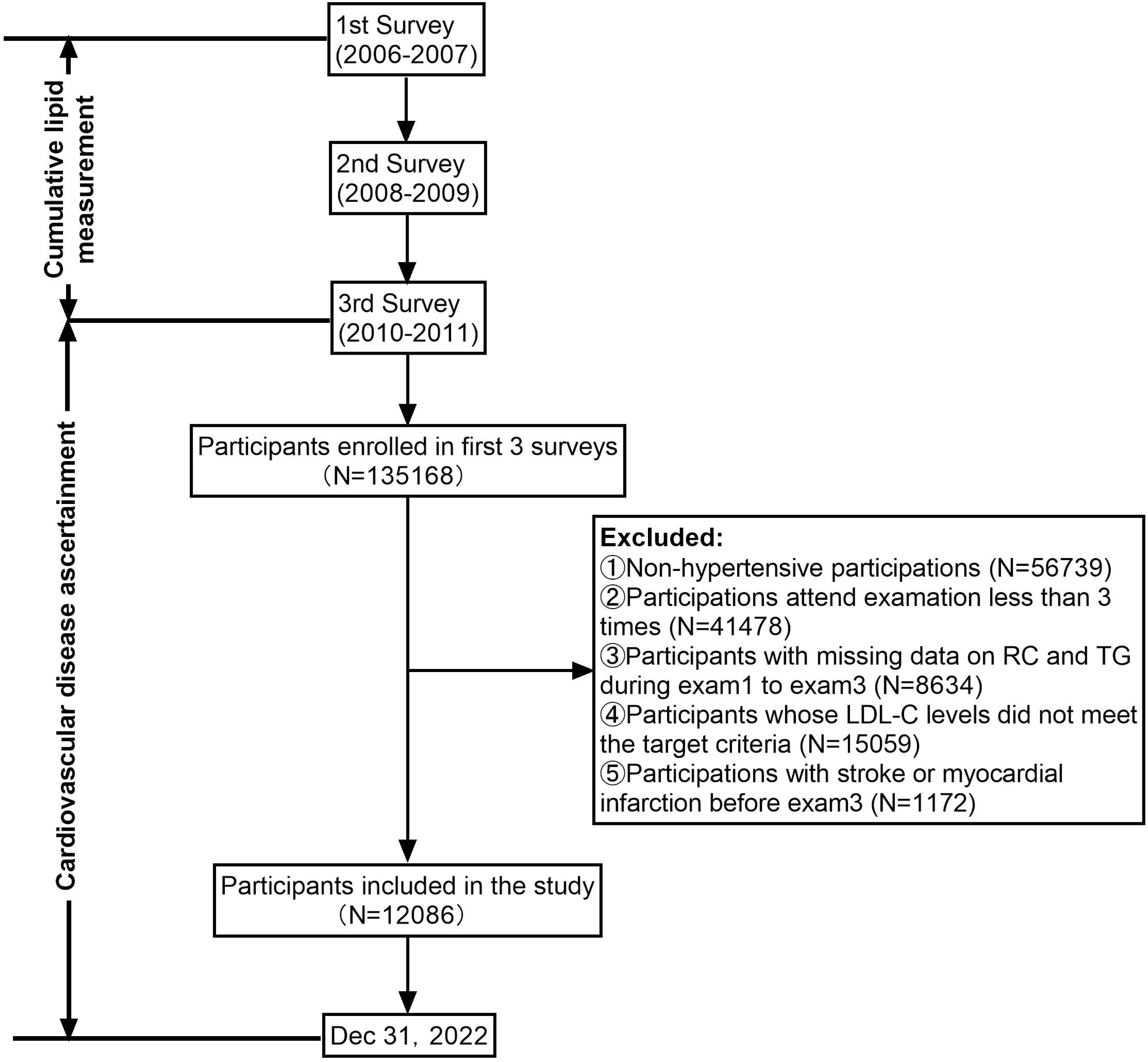
The flowchart of the study.

The study was approved by the ethics committee of Kailuan General Hospital (reference number: 2006-5) and performed in accordance with the Declaration of Helsinki. All participants provided written informed consent.

### Data collection

Data on related variables were collected through questionnaires, basic anthropometric measurements, and blood tests. Participants who had a history of smoking or currently smoked were defined as ever-smokers, and those who had a history of drinking or currently drank were defined as ever-drinkers. Physical activity was classified as never, occasional (1‒ 2 times/wk), or frequent (≥3 times/wk). Body mass index (BMI) was calculated as weight divided by height squared (kg/m^2^). Estimated glomerular filtration rate (eGFR) was calculated using the Chronic Kidney Disease Epidemiology Collaboration creatinine equation [13] and diabetes mellitus was defined as a fasting blood glucose level ≥7.00 mmol/L, taking antidiabetic agents, or a self-reported physician diagnosis of diabetes mellitus [14]. Blood pressure was measured in the left upper arm using a calibrated mercury sphygmomanometer with the participant in a sitting position. Concentrations of fasting blood glucose (FBG), C-reactive protein (CRP), total cholesterol (TC), TG, high-density lipoprotein cholesterol (HDL-C), and LDL-C were measured using an auto-analyzer (Hitachi 747; Hitachi, Tokyo, Japan) in the central laboratory of Kailuan hospital.

### Definition of cumulative RC and cumulative TG

RC was calculated by subtracting HDL-C and LDL-C from TC. The cumulative RC was computed as follows [15]: cumRC=[(RC_2006_+RC_2008_)/2×(Visit time_2008_ − Visit time_2006_)]+[(RC_2008_+RC_2010_)/2×(Visit time_2010_ − Visit time_2008_)]. In this equation, visit time_2006_, visit time_2008_, and visit time_2010_ were the dates of follow-up surveys during 2006, 2008, and 2010, respectively. Cumulative TG was calculated by the same way.

### Follow-up and primary outcomes

The outcome of this study was the composite of CVD events, including myocardial infarction (MI), ischemic stroke (IS), and hemorrhage stroke (HS). Participants were followed from the time of the physical examination in 2010 until the date of endpoint event, death or end of follow-up (31 December 2022).

MI was defined as elevated troponin T or troponin I with or without the following manifestations: ST-segment elevation on the electrocardiogram (ECG), or changes in myocardial ischemia on the ECG, or symptoms such as chest pain. Stroke was diagnosed according to the World Health Organization criteria on the basis of combined neurological signs and symptoms and imaging examinations, including computed tomography or magnetic resonance and was classified as IS and HS. Mortality information was collected from provincial vital statistics offices.

### Statistical Analysis

Continuous variables were described as means ± standard deviations (SD) or medians [interquartile ranges (IQR)] and compared using analysis of variance or the Kruskal-Wallis test according to distribution. Categorical variables were shown as n (%) and compared by the chi-square test. The cumulative incidence of end point events was estimated by Kaplan-Meier method and compared by log-rank test.

Cox proportional hazards model was used to explore the relationship between cumTG and cumRC and the risk of CVD in hypertensive patients with target LDL-C levels. The proportional hazards assumption was checked by using Schoenfeld residuals. Model 1 was adjusted for age and sex. Model 2 was further adjusted for BMI, SBP, LDL-C, FBG, eGFR, CRP, smoking status, drinking status, and physical activity. Model 3 was further adjusted for antihypertensive drugs, antidiabetic drugs, and lipid-lowering drugs. Sensitivity analyses were performed to assess robustness: (1) further adjusted for TG and RC in 2006; (2) further adjusted for TG and RC in 2010; (3) excluding participants taking lipid-lowering medications at baseline; (4) excluding participants with cancer at baseline; (5) excluding outcome events occurring within the first year of follow-up to minimize potential reverse causation; (6) We repeated our analyses using Fine-Gray models instead, accounting for the competing risk of death.

Stratified analyses by age (<60 years vs. ≥60 years), sex, BMI (<24 kg/m^2^ vs ≥24 kg/m^2^), FBG (<7 mmol/L vs. ≥7 mmol/L), and blood pressure control (yes or no) were performed to examine potential statistical interactions. We also examined the association of TG in 2010, RC in 2010, average TG, average RC, and their combined effect as secondary exposures with CVD risk.

All statistical analyses were conducted using SAS 9.4 (SAS Institute, Cray, NC, United States). Tests were two-sided, and a P-value < 0.05 was considered statistically significant.

## Results

### Baseline characteristics

A total of 12,086 participants (mean age was 55.44±11.56 years; 83.50% were men) were included and divided into four groups based on the median value of cumTG and cumRC: (1) low cumTG and low cumRC, (2) high cumTG and low cumRC, (3) low cumTG and high cumRC, and (4) high cumTG and high cumRC. Compared with the groups of low cumTG and low cumRC, participants with high cumTG and high cumRC were more likely to be female, smokers and drinkers, to have higher BMI, blood pressure, TC, TG, CRP, eGFR, and FBG, a higher prevalence of diabetes mellitus, and a higher proportion of taking antihypertensive, antidiabetic, and lipid-lowering drugs, but to have lower levels of LDL-C and HDL-C, and less likely to be physically active (Table 1).

**Table 1.** Baseline characteristics of participants according to categories of low and high cumTG and cumRC

|  | Low cumTG and<br>low cumRC<br>(n=4001) | High cumTG and<br>low cumRC<br>(n=2054) | Low cumTG and<br>high cumRC<br>(n=2034) | High cumTG and<br>high cumRC<br>(n=3997) | *P* value |
| --- | --- | --- | --- | --- | --- |
| Age (mean±SD, years) | 55.26±12.35 | 53.36±11.72 | 58.30±10.53 | 55.24±10.85 | <0.01 |
| Man [n(%)] | 3347 (83.65) | 1766 (85.98) | 1675 (82.35) | 3298 (82.51) | <0.01 |
| Ever-smokers [n(%)] | 1415 (35.37) | 851 (41.43) | 732 (35.99) | 1723 (43.11) | <0.01 |
| Ever-drinkers [n(%)] | 1371 (34.27) | 814 (39.63) | 628 (30.88) | 1569 (39.25) | <0.01 |
| Physical activity [n(%)] |  |  |  |  | <0.01 |
| Never | 1209 (30.22) | 680 (33.11) | 824 (40.51) | 1552 (38.83) |  |
| Occasionally | 2232 (55.79) | 1105 (53.80) | 952 (46.80) | 1936 (48.44) |  |
| Frequently | 560 (14.00) | 269 (13.10) | 258 (12.68) | 509 (12.73) |  |
| Diebets mellitus [n(%)] | 487 (12.17) | 407 (19.81) | 306 (15.04) | 975 (24.39) | <0.01 |
| BMI (mean±SD, kg/m<sup>2</sup>) | 24.62±3.41 | 26.20±3.30 | 24.89±3.24 | 26.47±3.25 | <0.01 |
| SBP (mean±SD, mm Hg) | 137.25±18.13 | 138.56±17.52 | 137.65±16.89 | 139.76±17.49 | <0.01 |
| DBP (mean±SD, mm Hg) | 87.45±10.31 | 88.97±10.22 | 87.42±9.32 | 89.37±10.02 | <0.01 |
| FBG (mean±SD, mmol/L) | 5.54±1.37 | 5.76±1.62 | 5.65±1.50 | 5.99±1.83 | <0.01 |
| CRP [M(IQR), mg/L] | 1.14 (2.14) | 1.20 (2.28) | 1.00 (2.90) | 1.44 (2.77) | <0.01 |
| RC [M(IQR), mmol/L] | 0.45 (0.52) | 0.79 (0.66) | 1.52 (1.36) | 1.65 (1.38) | <0.01 |
| TC (mean±SD, mmol/L) | 4.28±0.62 | 4.41±0.62 | 4.92±0.79 | 5.10±1.00 | <0.01 |
| TG [M(IQR), mmol/L] | 0.88 (0.50) | 1.67 (1.12) | 1.00 (0.50) | 2.26 (1.89) | <0.01 |
| LDL-C (mean±SD, mmol/L) | 2.06±0.41 | 2.12±0.39 | 1.81±0.55 | 1.92±0.51 | <0.01 |
| HDL-C (mean±SD, mmol/L) | 1.61±0.44 | 1.43±0.40 | 1.56±0.44 | 1.36±0.39 | <0.01 |
| eGFR (mean±SD,<br>mL/min/1.73m<sup>2</sup>) | 87.93±18.69 | 88.98±17.85 | 90.93±15.18 | 91.00±17.17 | <0.01 |
| Antidiabetic drugs [n(%)] | 178 (4.44) | 142 (6.91) | 150 (7.37) | 374 (9.36) | <0.01 |
| Antihypertensive drugs [n(%)] | 1017 (25.42) | 713 (34.71) | 599 (29.45) | 1661 (41.56) | <0.01 |
| Cholesterol-lowering drugs<br>[n(%)] | 43 (1.07) | 44 (2.14) | 42 (2.06) | 179 (4.48) | <0.01 |

TG indicates triglyceride; RC, remnant cholesterol; BMI, body mass index;SBP, systolic blood pressure ;DBP, diastolic blood pressure; FBG, fasting blood glucose; CRP, C-reactive protein; TC, total cholesterol; LDL-C, low-density lipoprotein cholesterol; HDL-C, high-density lipoprotein cholesterol; eGFR, estimated glomerular filtration rate.

### Joint associations of cumTG and cumRC with the risk of CVD

During a median follow-up period of 11.95 years, 1,392 participants experienced CVD events (including 423 cases of MI, 891 cases of IS and 138 cases of HS). The Kaplan-Meier curves shows that the high cumTG/high cumRC group had the highest risk of CVD compared to the low cumTG/low cumRC group, followed by the low cumTG/high cumRC group and the high cumTG/low cumRC group (log-rank *P*< 0.01, Figure 2). Compared with the low cumTG/low cumRC group, participants in the high cumTG/high cumRC group were associated with a higher risk of CVD (HR 1.43, 95%CI 1.24‒1.64), MI (HR 1.81, 95%CI 1.40‒2.34), and IS (HR 1.39, 95%CI 1.17‒1.65), but not with the risk of HS (HR 0.80, 95%CI 0.51‒1.25). Similar relationships were observed in the low cumTG/high cumRC group (Figure 3).

**Figure 2.**
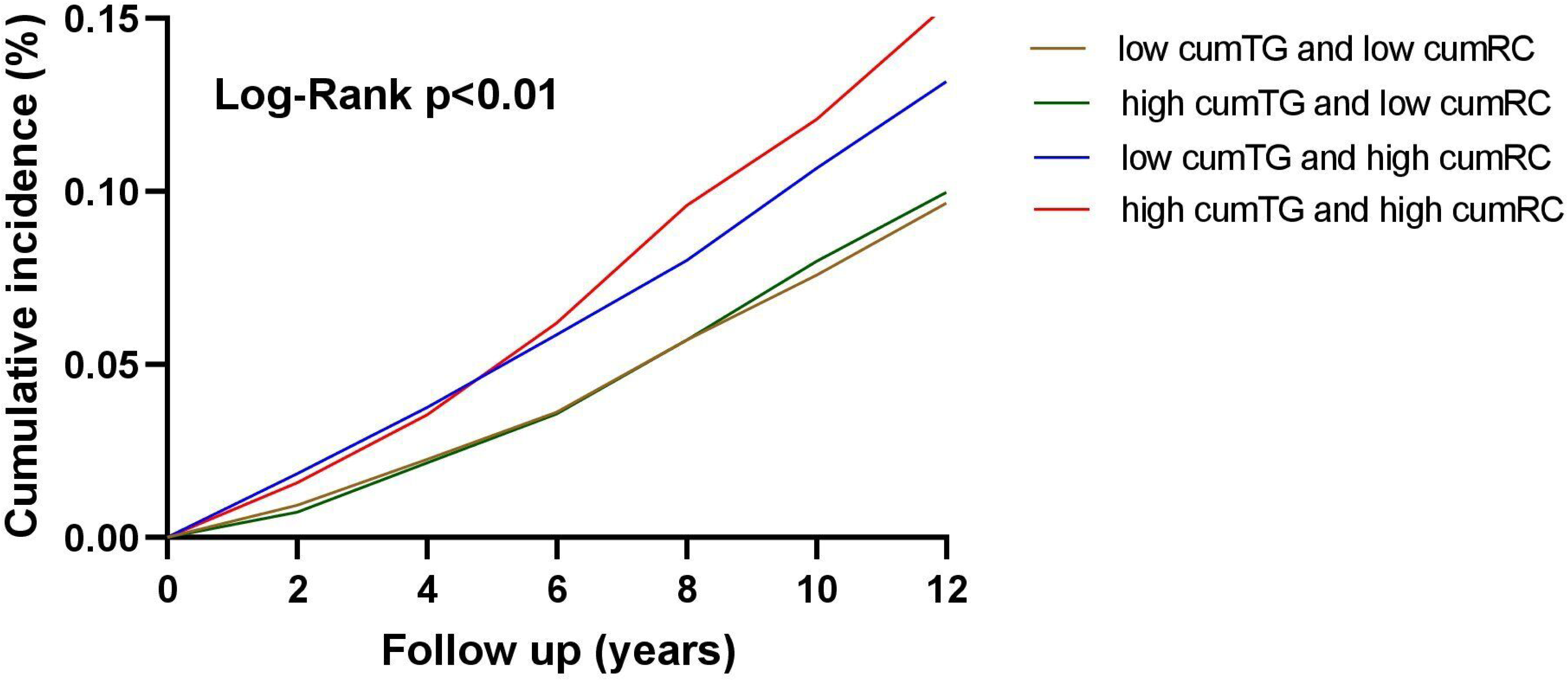
Kaplan-Meier estimates of incidence of CVD during follow-up according to cumTG and cumRC groups.

**Figure 3.**
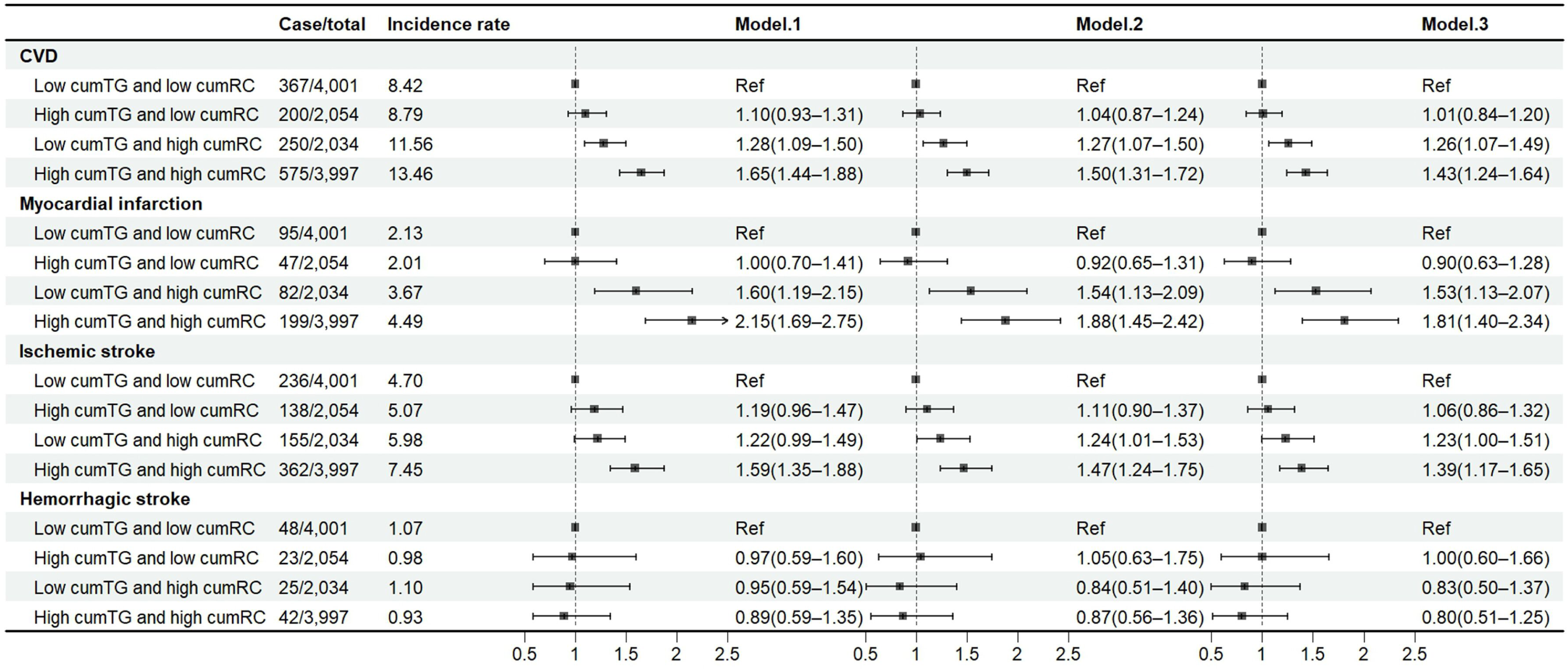
Association between combined effect of cumTG and cumRC and the risk of CVD.

### Separate associations of cumTG and cumRC with the risk of CVD

The cumulative incidences of CVD increased across the Q1‒Q4 quartile groups by cumTG and cumRC and significantly differed (both log-rank *P*<0.01, Figure 4‒5). Participants in quartile 4 of cumTG (HR 1.35, 95% CI 1.16‒1.59) or cumRC (HR 1.58, 95% CI, 1.35‒1.86) had a higher risk of CVD compared with those in quartile 1 (Figure 6). Moreover, cumTG and cumRC were associated with a 2% and 7% higher risk of CVD per every 1 mmol/L increase, respectively (Table S1).

**Figure 4.**
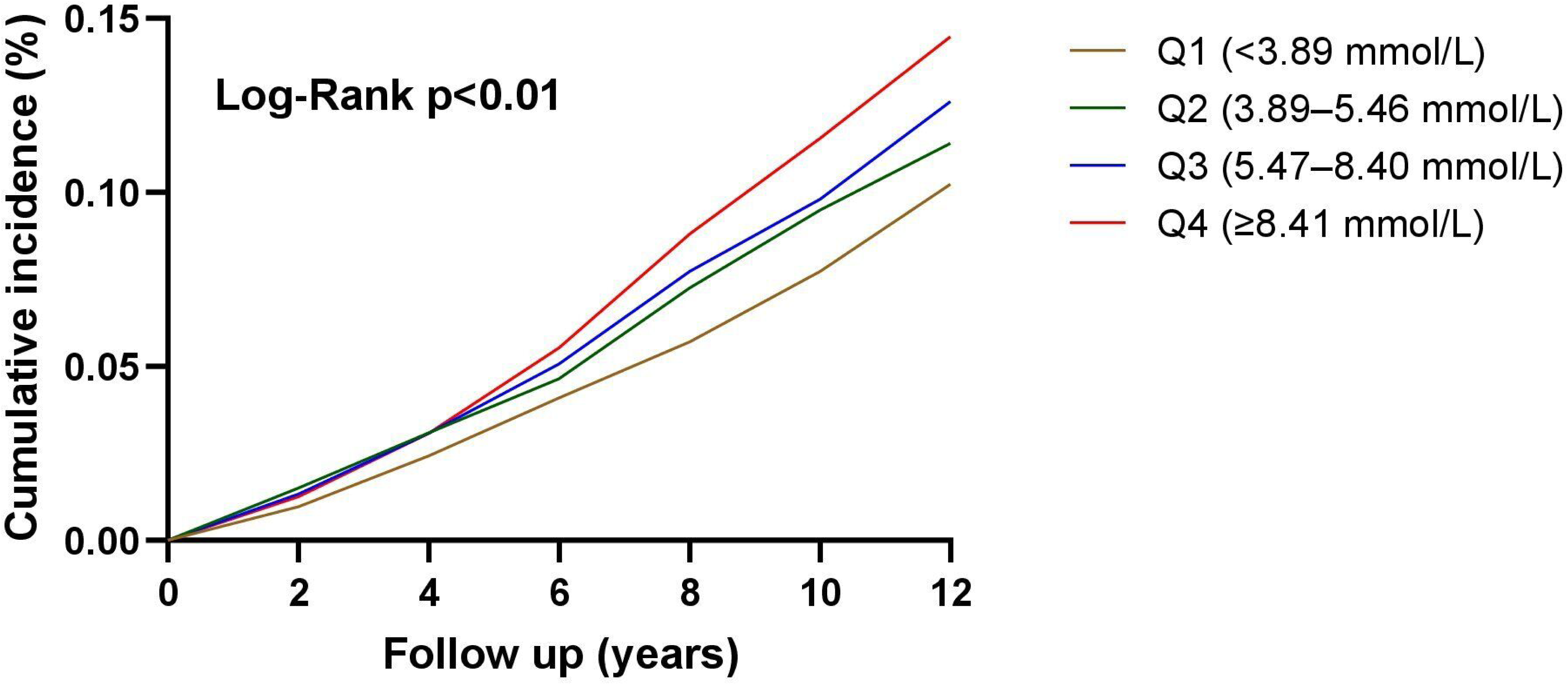
Kaplan-Meier estimates of incidence of CVD during follow-up according to quartiles of cumTG.

**Figure 5.**
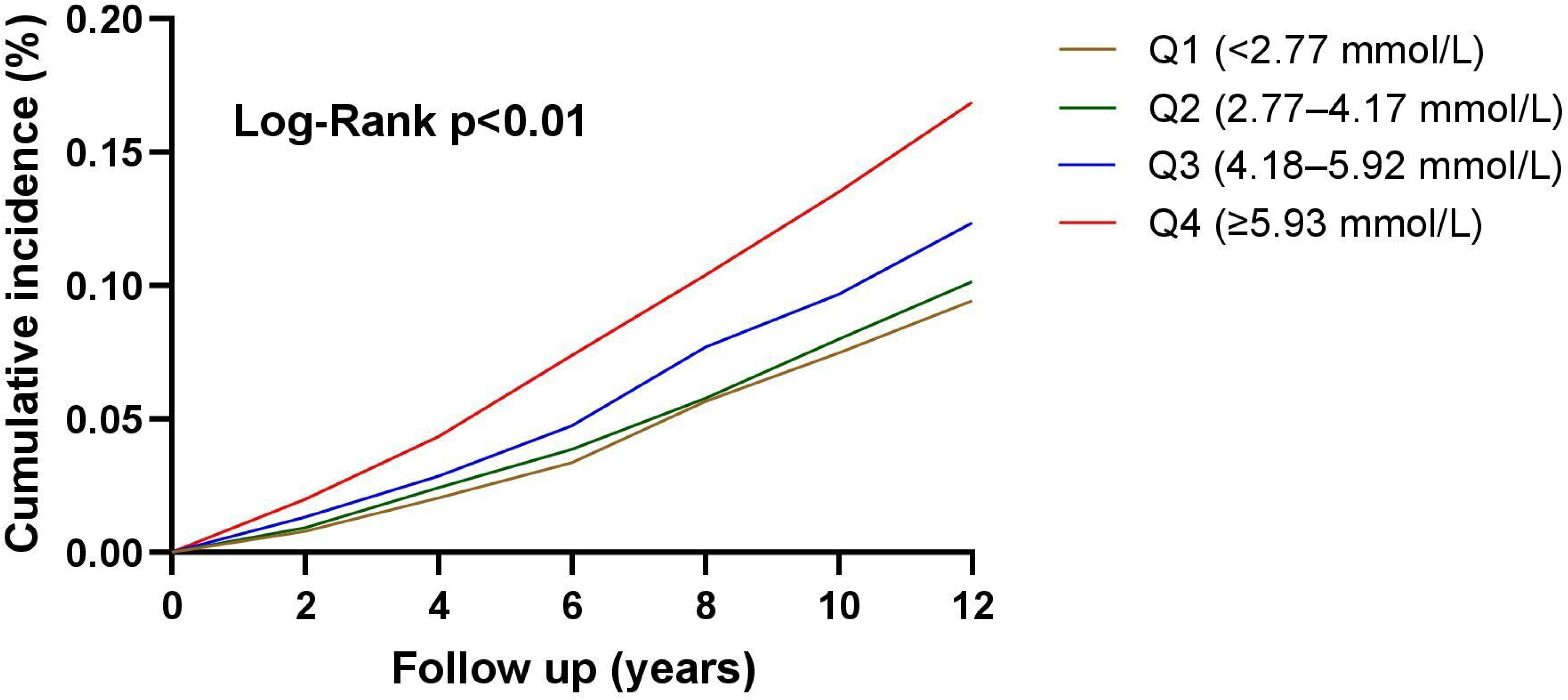
Kaplan-Meier estimates of incidence of CVD during follow-up according to quartiles of cumRC.

**Figure 6.**
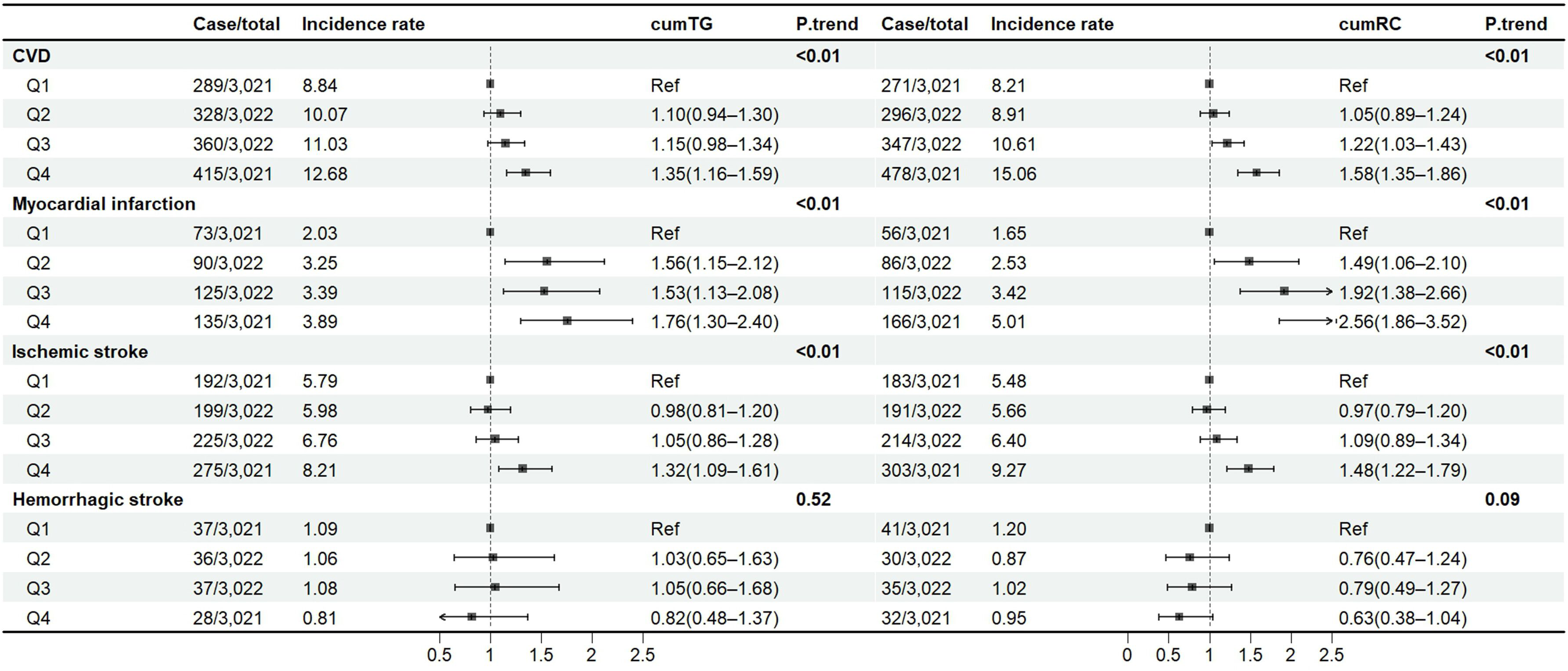
Association between separate effect of cumTG and cumRC and the risk of CVD. **Figure legend:** Model was adjusted for age, gender, BMI, SBP, LDL-C, eGFR, FBG, CRP, drinking status, smoking status, physical activity, antihypertensive medication, lipid-lowering medication, and antidiabetic medication.

### Sensitivity and stratified analyses

The outcomes remained consistent when we further adjusted for TG and RC in 2006, TG and RC in 2010, excluded participants using lipid-lowering drugs, excluded participants with cancer at baseline, and excluded cases of CVD that occurred within the first year of follow-up. Additionally, to account for the competing risk of mortality, we conducted Fine-Gray model and the findings remained stable (Table S2‒S4).

Subgroup analyses showed that there was no interaction between cumTG-cumRC groups and stratified groups including age, BMI, FBG, and blood pressure control (all P for interaction > 0.05), except for sex (P for interaction=0.03). In addition, the association of cumTG with CVD appeared to be stronger in man (P for interaction=0.04) (Table S5‒S7).

### Secondary analysis

In the secondary analyses, the highest quartile of TG (in 2010 and average) and RC (in 2010 and average) were associated with a higher risk of CVD, compared with the lowest quartile group (Table S8). When considering the combined effects of TG and RC, participants with high TG and high RC (HR 1.35, 95% CI 1.17‒1.55) or low TG and high RC (HR 1.29, 95%CI 1.09‒1.52) had a higher CVD risk than those with low TG and low RC (Table S9‒ S10).

## Discussion

In the present study, we found that cumulative TG and RC burdens were independently associated with CVD risk in hypertensive patients with target LDL-C levels. High levels of cumRC were more associated with increased CVD risk compared with cumTG, indicating that RC is the primary residual risk factor for CVD beyond LDL-C and TG. Lipid-lowering interventions should not only focus on lowering TG levels but also on maintaining low RC levels.

A meta-analysis including 29 studies with 257,387 participants found that RC levels were associated with ASCVD risk both in primary and secondary prevention [16]. A single measurement is insufficient to reflect the longitudinal effect of RC on the risk of CVD, which enhances the measurement of cumulative exposure burden. A 30-year population-based cohort study concluded that higher cumulative exposure to RC increased the risk of CVD events independent of LDL-C levels [17]. Another Kailuan study found that high cumulative RC exposure significantly associated with a higher risk of CVD in individuals with hypertension than those without [18]. Based on prior studies, our study further extended to the hypertensive patients who have reached the target LDL-C levels. To the best of our knowledge, the prospective cohort study explored the separate and joint associations of TG and RC accumulation with CVD in hypertensive patients with target LDL-C levels. We found that the risk of CVD was 1.26 higher in the high cumRC and low cumTG group and 1.43 higher in high cumRC and high cumTG group. A meta-analysis including 302,430 participants without vascular disease from 68 long-term prospective studies found that the HR for coronary heart disease with TG reduced from 1.37 (95% CI, 1.31‒1.42) to 0.99 (95% CI, 0.94‒1.05) after further adjustment for HDL-C and non-HDL-C [19], indicating that TG’s contribution to CVD risk is mediated by these lipoproteins rather than being an independent factor. In contrast, RC was independently associated with cardiovascular outcomes, as demonstrated by a Mendelian randomization study [20]. Different pharmacologic therapies have been explore to determine if a reduction in TG and RC translates to a reduction in CVD events, with randomized controlled trials (RCTs) resulting in heterogeneous findings [21–23]. Whether further lowering RC is effective in hypertensive patients with already optimized LDL levels requires further research.

Several potential mechanisms may explain the association between cumulative RC exposure and CVD outcomes. First, cholesterol could accumulate in intimal foam cells, not TG, which makes the RC become the atherogenic content of TRLs [24]. RC holding four times more cholesterol per particle than LDL-C and are sufficiently small (< 70 nm diameter) to penetrate the arterial intima and retain in the sub-endothelium after lipolysis [25]. In addition, RC could be taken up by macrophages directly leading to foam cell formation without oxidative modification [26], which contribute to cholesterol deposition, prothrombotic initiation and ASCVD-related events progression. Second, RC levels are highly associated with low-grade inflammation, which results in vascular injury and atheroma [27]. Mendelian randomization suggests a causal association of elevated RC with inflammation and ischemic heart disease [28], indicating that inflammation of atherosclerosis is driven by elevated RC levels. Our research also determined that cumRC remains an independent CVD risk factor after adjusting CRP. Finally, increased levels of RC may be associated with increased insulin resistance [29]. For hypertensive patients, it is essential to control RC levels in addition to LDL-C and TG. Prioritizing the development of targeted therapies to reduce RC may be crucial for mitigating residual lipid-associated CVD risk.

This study has several strengths. We investigated the combined effect of TG and RC with the risk of CVD in hypertensive patients with target LDL-C level based on a large prospective cohort study. By measuring the cumulative exposures, our study increases the reliability of the findings than those relied on a single measurement. This study also has some limitations. First, RC levels were not directly measured but obtained by calculation, which may cause bias. However, it has been shown that calculated RC concentrations correlate well with direct measurements and the calculated RC is widely used in many population studies [30]. Second, there may exist residual confounding factors that were not identified even after a range of sensitivity analyses. Third, our findings were based on Northern Chinese coal miners, so the results may not be generalizable to other populations. Fourth, due to limited available data, lipoprotein(a), apolipoprotein B -containing lipoproteins, as a potential new target for lipid intervention in recent years, was not analysed in our study. Finally, our findings were based on observational data, and additional research is warranted to explore the pathophysiological mechanisms underlying the effect of cumulative RC exposure on CVD.

## Conclusion

Our findings provide support that the cumulative RC burden in hypertensive patients with target LDL-C was associated with the risk of CVD beyond cumTG. The findings suggest that maintaining optimal RC levels, not TG alone, may further reduce the risk of cardiovascular events when LDL-C is within the target range.

## Data Availability

The data that support the findings of this study are available from [third party name] but restrictions apply to the availability of these data, which were used under license for the current study, and so are not publicly available. Data are however available from the corresponding author upon reasonable request and with permission of the corresponding author

## Acknowledgements

We sincerely thank all the survey teams of the Kailuan Study Group for their contribution and the study participants who contributed their information.

## Sources of Funding

None.

## Disclosures

The authors declare no conflicts of interest.

## Supplemental Material

**Table S1.** Hazard ratios (95% CI) for CVD according to cumRC and cumTG (as continuous variables).

**Table S2.** Sensitivity analysis of CVD according to four cumTG/cumRC groups in hypertensive patients with target LDL-C

**Table S3.** Sensitivity analysis of CVD according to cumTG in hypertensive patients with target LDL-C

**Table S4.** Sensitivity analysis of CVD according to cumRC in hypertensive patients with target LDL-C

**Table S5.** Stratified analyses for the association of four cumTG/cumRC groups with CVD

**Table S6.** Stratified analyses for the association of cumulative TG with CVD **Table S7.** Stratified analyses for the association of cumulative RC with CVD **Table S8.** Secondary analyses for the association of TG and RC with CVD

**Table S9.** Secondary analyses for the combined effect of RC and TG levels in 2010 with CVD

**Table S10.** Secondary analyses for the combined effect of average RC and average TG with CVD

## Notes

### Competing Interest Statement

The authors have declared no competing interest.

### Author Declarations

The study protocol was approved by the ethics committee of the Kailuan General Hospital (approval number: 2006-5), and all participants signed informed consent prior to study inclusion.

